# Determinants of in-hospital mortality in COVID-19; a prospective cohort study from Pakistan

**DOI:** 10.1101/2020.12.28.20248920

**Authors:** Samreen Sarfaraz, Quratulain Shaikh, Syed Ghazanfar Saleem, Anum Rahim, Fivzia Farooq Herekar, Samina Junejo, Aneela Hussain

## Abstract

A prospective cohort study was conducted at the Indus Hospital Karachi, Pakistan between March and June 2020 to describe the determinants of mortality among hospitalized COVID-19 patients. 186 adult patients were enrolled and all-cause mortality was found to be 36% (67/186). Those who died were older and more likely to be males (p<0.05). Temperature and respiratory rate were higher among non-survivors while Oxygen saturation was lower (p<0.05). Serum CRP, D-dimer and IL-6 were higher while SpO2 was lower on admission among non-survivors (p<0.05). Non-survivors had higher SOFA and CURB-65 scores while thrombocytopenia, lymphopenia and severe ARDS was more prevalent among them (p<0.05). Use of non-invasive ventilation in emergency room, ICU admission and invasive ventilation were associated with mortality in our cohort (p<0.05). Length of hospital stay and days of intubation were longer in non-survivors (p<0.05). Use of azithromycin, hydroxychloroquine, steroids, tocilizumab, antibiotics, IVIG or anticoagulation showed no mortality benefit (p>0.05). Multivariable logistic regression showed that age > 60 years, oxygen saturation <93% on admission, pro-calcitonin > 2 ng/ml, unit rise in temperature and SOFA score, ICU admission and sepsis during hospital stay were associated with higher odds of mortality. Larger prospective studies are needed to further strengthen these findings.

**Key Findings:**

1. Age greater than 60 years is associated with in-hospital mortality among COVID-19 patients
2. Oxygen saturation less than 93% and ICU admission are associated with higher odds of mortality
3. Inflammatory markers including CRP, Ferritin and IL-6 were significantly higher among non-survivors
4. Serum pro-calcitonin greater than 2 ng/ml and sepsis during hospital stay are associated with higher odds of mortality among COVID-19 patients

## Introduction

In December 2019 a highly transmissible respiratory illness caused by a novel corona virus, later named severe acute respiratory syndrome coronavirus 2(SARS-Cov-2), originated in Wuhan, China and caused a pandemic by its rapid spread [1]. It has resulted in 79,057,616 infections and 1,737,751 deaths globally as of 24^th^ December 2020. Pakistan ranks 28^th^ among the list of high burden countries with 465,070 confirmed infections and 9,668 deaths as of December 24^th^ which is much lower compared to its immediate neighbours [2]. Our mortality rate of 2% [3] is comparable to the India (1.45%) but lower than Iran (4.68%) and several European countries including UK (3.43%) and Italy (3.52%)[2]. The reasons for this difference in fatality is largely unknown but a multifactorial combination of viral immunogenicity, genetic makeup of the host, demography and seasonal variation may play a role in this [4]. Sind province recorded the country’s first case on 26^th^ February 2020 and since then has received 44% of the country’s COVID-19 confirmed cases with the largest city Karachi being worst hit [3]. The peak of infection in the first wave reached in mid-June when on average 7000 new infections were recorded in a day and the maximum number of deaths recorded were 153 on 20^th^ June 2020. Major hospitals in all big cities were overwhelmed straining the health infra-structure. The Indus Hospital, Karachi emerged as a front liner with a dedicated isolation unit providing free of cost treatment to the sick COVID-19 patients requiring hospitalization. The hospital worked with the government of Sind as a referral and diagnostic center. The experience was challenging for our physicians and allied health staff. We planned to estimate the in-hospital mortality of COVID-19 in Pakistan and study it’s determinants in our patients. Till date there is insubstantial published data on in-hospital mortality from Pakistan. As Pakistan has already entered the second wave now, this data will help in risk stratification and management of COVID-19 patients.

## Methods

This prospective cohort study was conducted at the Indus Hospital’s COVID isolation unit. The Indus Hospital (TIH) Karachi is a 300 bedded tertiary care hospital set up with public private partnership which provides free of cost services to the people. The COVID unit was initially a 20 bedded dedicated COVID facility established in March 2020, later expanded to 56 beds. All COVID-19 (Nasopharyngeal PCR positive) patients older than 18 years admitted between 19th March and 7th June 2020 were included. The study was approved by the institutional IRB under IRD_IRB_2020_04_002. Demographic information, clinical presentation, laboratory abnormalities including inflammatory markers and imaging results were recorded. Patients were categorized as per WHO definitions into asymptomatic (COVID-19 PCR positive but with no clinical manifestation attributable to COVID-19), mild (symptomatic without evidence of pneumonia), moderate (with clinical signs of pneumonia and oxygen saturation ≥ 90% on room air), severe (signs of pneumonia with respiratory rate ≥ 30 breaths/min or SpO2 < 90% on room air) and critical (development of Acute Respiratory Distress Syndrome, septic shock or multi organ dysfunction)[5]. Usually patients who required ventilator and medical support were admitted. Few patients needed admission due to other reasons like emergency surgery, pyelonephritis and routine dialysis and these were also included in the cohort. Patients were enrolled into the study as soon as they were admitted in the COVID unit. Patients were followed till death or discharge from hospital. The primary outcome was all-cause in-hospital mortality while we also compared length of stay, occurrence of in-hospital complications, use of inotropic support and of mechanical ventilation among survivors and non survivors. Data was recorded on a REDCap database and analyzed on Stata14. Continuous variables were expressed as mean (SD) or median (IQR) as appropriate. Categorical variables were expressed as number (%). Student’s t-test or Mann Whitney U test were used to compare continuous data. Chi square or Fischer’s Exact test was used to compare categorical data. Univariable binary logistic regression was used to model mortality against the predictors. All variables with p-value <0.10 were considered for multivariable binary logistic regression model in order of significance. Odds ratios and 95% confidence intervals were reported.

## Results

The emergency department of Indus hospital, Karachi received 11,855 suspected COVID-19 patients out of which 3,851 tested positive for COVID-19 PCR (positivity rate 32.5%) from 19th March to 7th June 2020 (Fig 1). The median age of those who tested positive for COVID-19 PCR was 35 (IQR=26-50) years. The teleconsultation service set up for COVID-19 at TIH, managed most (3,422; 88%) of these asymptomatic to mild spectrum patients at home through a robust algorithm based system of symptomatic treatment, follow up and counselling. Among the rest 384 (9.9%) needed admission but due to limited bed availability in the isolation unit, only 193 (50%) were admitted while the rest were referred to other isolation units in the city. Seven patients who were admitted at TIH were less than 15 years old hence removed from this analysis. Eventually, 186 participants were included in the cohort. The all-cause in-hospital mortality was 36% (67/186). Those who died had a median age 61.4 years (Table 1) compared to those who survived 51.3 years (p<0.001). Males were more likely to die (77.6% in non-survivors compared to 76.5%, p<0.05) in the study cohort.

**Table 1:** Clinical characteristics of study participants by mortality.

|  | **Non-Survivors**<br>**n(%)** | **Survivors**<br>**n(%)** | **All patients** | **p-value** |
| --- | --- | --- | --- | --- |
| **Age<sup>^^</sup> (years)** | 61.4(54.1-69.1) | 51.3(42.1-62.2) | 57(45.3-65.1) | 0.000<sup>**M</sup> |
| **Age (years)** |  |  |  |  |
| ≤60 | 24(35.8) | 74(62.2) | 98(52.7) | 0.000<sup>**P</sup> |
| >60 | 43(64.2) | 45(37.8) | 88(47.3) |  |
| **Gender** |  |  |  |  |
| Male | 52(77.6) | 91(76.5) | 148(76.9) | 0.859<sup>P</sup> |
| Female | 15(22.4) | 28(23.5) | 43(23.1) |  |
| **Symptoms** |  |  |  |  |
| Fever | 54(81.8) | 91(77.8) | 145(79.2) | 0.155<sup>MC</sup> |
| Cough | 40(60.6) | 67(57.3) | 107(58.5) |  |
| SOB | 57(86.4) | 88(75.2) | 145(79.2) |  |
| Runny nose | 1(1.5) | 4(3.4) | 5(2.7) |  |
| Sore Throat | 3(4.5) | 4(3.4) | 7(3.8) |  |
| Chest Pain | 5(7.6) | 4(3.4) | 9(4.9) |  |
| Fatigue | 8(12.1) | 7(6.0) | 15(8.2) |  |
| Diarrhea | 4(6.1) | 10(8.5) | 14(7.7) |  |
| Vomiting | 2(3.0) | 14(12.0) | 16(8.7) |  |
| Others | 13(19.7) | 33(28.2) | 46(25.1) |  |
| **Total Duration of symptoms<sup>^</sup>(days)** | 6(4-7) | 7(4-10) | 6(4-7) | 0.121<sup>M</sup> |
| **Comorbid conditions** |  |  |  |  |
| None | 13(19.4) | 47(39.5)b | 60(32.3) | 0.007<sup>*M</sup><br><sup>C</sup> |
| Hypertension | 35(52.2) | 43(36.1) | 78(41.9) |  |
| Diabetes | 35(52.2) | 44(37.0) | 79(42.5) |  |
| Liver Disease | 1(1.5) | 0 | 1(0.5) |  |
| Lung Disease | 4(6.0) | 3(2.5) | 7(3.8) |  |
| Renal Disease | 6(9.0) | 11(9.2) | 17(9.1) |  |
| Heart Disease | 7(10.4) | 11(9.2) | 18(9.7) |  |
| Other | 18(26.9) | 41(34.5) | 59(31.7) |  |
| **Number of comorbid conditions** |  |  |  |  |
| ≤2 | 43(79.6) | 50(69.4) | 93(73.8) | 0.198<sup>P</sup> |
| 3-5 | 11(20.4) | 22(30.6) | 33(26.2) |  |
| **Clinical Signs at presentation** |  |  |  |  |
| **Systolic Blood pressure<sup>^</sup> (mm/Hg)** | 144 (123-159) | 134 (120-149) | 138.5(121-153) | 0.023<sup>*M</sup> |
| **Diastolic Blood pressure<sup>^</sup> (mm/Hg)** | 80(69-95) | 79(72-89) | 79.5(70-90) | 0.982<sup>M</sup> |
| **Pulse<sup>^^</sup>/minute** | 104.6 ± 22.4 | 99.9 ± 19.9 | 101.6 ± 20.9 | 0.141<sup>T</sup> |
| **Respiratory Rate<sup>^</sup>/m** | 32(26-38) | 28(22-32) | 28(24-36) | 0.000<sup>**M</sup> |
| **Temperature<sup>^</sup> °F** | 98.6(98-99.1) | 98.3(98-98.6) | 98.6(98-98.6) | 0.003<sup>*D</sup> |
| **Oxygen saturation<sup>^</sup> (%)** | 86(69-90) | 93(86-96) | 90(82-95) | 0.000<sup>**M</sup> |
| **Oxygen Saturation (%)** |  |  |  |  |
| ≥93 | 13(19.4) | 62(52.1) | 75 (40.3) | 0.000<sup>*P</sup> |
| <93 | 54(80.6) | 57(47.9) | 111 (59.7) |  |
| **GCS** | 15(15-15) | 15(15-15) | 15(15-15) | 0.452<sup>M</sup> |
| **Disease Severity** |  |  |  |  |
| Asymptomatic | 0 | 2(1.3) | 2(1.1) | 0.000<sup>**f</sup> |
| Mild | 0 | 13(10.9) | 13(7.0) |  |
| Moderate | 0 | 18(15.1) | 19(9.7) |  |
| Severe | 3(4.5) | 13(10.9) | 16(8.6) |  |
| Critical | 64(95.5) | 73(61.3) | 136(73.7) |  |
| <sup>^</sup>Median (IQR), <sup>^^</sup>Mean±SD, *P-value<0.05, **P-value<0.0001, T: Independent sample T test, M: Mann Witney U test, P: Pearson Chi-Square, f: Fisher's Exact Test, MC: Chi-square for multiple Responses |  |  |  |  |
| <sup>^</sup>Median (IQR), <sup>^^</sup>Mean±SD, *P-value<0.05, **P-value<0.0001, T: Independent sample T test, M: Mann Witney U test, P: Pearson Chi-Square, f: Fisher's Exact Test, MC: Chi-square for multiple Responses |  |  |  |  |

**Figure 1:**
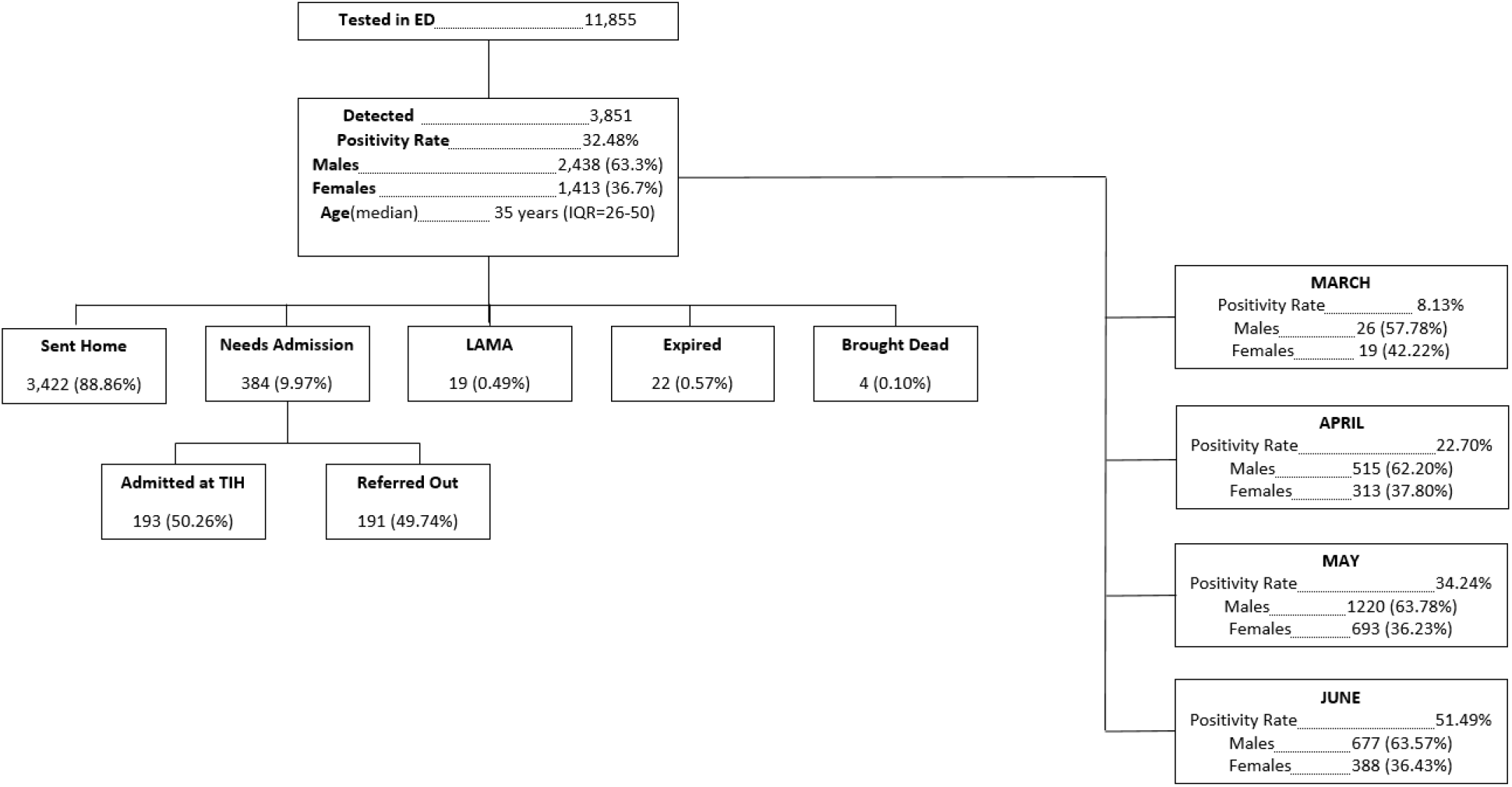
Flow and outcomes of COVID-19 suspects at the Indus Hospital Emergency (19^th^ March-7^th^ June 2020)

### Clinical presentation

Fever, shortness of breath and cough were three most common symptoms in both groups while fatigue was common in non-survivors and vomiting was seen in survivors (p>0.05). Hypertension or diabetes were the most prevalent comorbid conditions (52% vs 36% and 52% vs 37% among non-survivors compared to survivors respectively, p=0.007). Blood pressure and heart rate were similar in the groups but respiratory rate was 32/min in non-survivors while 28/min in survivors, p<0.001). Median peripheral oxygen saturation was 86% among non-survivors compared to 93% among survivors (p<0.00). Temperature was also higher among non-survivors (p=0.003). Admitted patients were usually critical (95% among non-survivors vs 61% among survivors, p<0.001). Majority of non-survivors required non-invasive ventilation in the emergency room (56.7 % vs 42%, p<.0001)

### Baseline laboratory parameters and Clinical severity scores

At admission, the WBC count and absolute neutrophil count was higher among non-survivors (12.5 vs 9.9 × 109 /L, p=0.021) and (10.4 vs 7.9 × 109/L, p<0.001) respectively (Table 2). While absolute lymphocyte count and platelet count was lower among non survivors (0.9 vs 1.1 × 109 /L, p=0.009) and (198 vs 234 X109/L, p=0.045). Neutrophil/Lymphocyte ratio (NLR) was higher among non-survivors (10.7 vs 6.7, p<0.001).

**Table 2:** Baseline laboratory parameters and Clinical severity scores by mortality.

|  |  |  |  |  |
| --- | --- | --- | --- | --- |
| **Hemoglobin<sup>^</sup>(gm/dl)** | 12.7(11.4-14.3) | 13.1(11.6-14.1) | 13(11.6-14.1) | 0.894<sup>M</sup> |
| **Total WBC Count<sup>^</sup> (x10E9/L)** | 12.5(8.5-15.5) | 9.9(6.9-13.9) | 10.8(7.1-14.7) | 0.021<sup>*M</sup> |
| **Neutrophils<sup>^</sup> (%)** | 86.3(81-91.2) | 80.6(75-87.5) | 83 (76.5-89) | 0.000<sup>**M</sup> |
| **Lymphocytes<sup>^</sup> (%)** | 8.1(4.9-12.6) | 12.2(7.8-18.1) | 10.7(6.9-16.1) | 0.000<sup>**M</sup> |
| **Eosinophils<sup>^</sup> (%)** | 0(0-0.1) | 0.1(0-0.3) | 0(0-0.2) | 0.000<sup>**M</sup> |
| **Platelet Count<sup>^</sup> (x10E9/L)** | 198(147-296) | 234.5(187-326) | 232(169-323) | 0.045<sup>*M</sup> |
| **ANC<sup>^</sup> (x10E9/L)** | 10.4(7.6-14.0) | 7.9(5.3-11.6) | 8.9(5.7-12.6) | 0.006<sup>*M</sup> |
| **ALC<sup>^</sup> (x10E9/L)** | 0.9(0.5-1.3) | 1.1(0.7-1.7) | 1.0(0.7-1.7) | 0.009<sup>*M</sup> |
| **ALC** |  |  |  |  |
| ≤1 | 39(58.2) | 52(44.1) | 91(49.2) | 0.064<sup>*P</sup> |
| >1 | 28(41.8) | 66(55.9) | 94(50.8) |  |
| **NLR<sup>^</sup>** | 10.7(6.2-18.8) | 6.7(4.2-11.2) | 8.0(4.7-13.0) | 0.000<sup>**M</sup> |
| **NLR** |  |  |  |  |
| <5 | 9(13.4) | 43(36.1) | 52(28.0) | 0.001<sup>*P</sup> |
| ≥5 | 58(86.6) | 76(63.9) | 134(72.0) |  |
| **Arterial pH<sup>^</sup>** | 7.4(7.4-7.5) | 7.4(7.4-7.5) | 7.4(7.4-7.5) | 0.421<sup>M</sup> |
| **PCO2 (mmHg)<sup>^</sup>** | 31.3(27.2-35.8) | 31.9(28.5-34.7) | 31.6(27.7-34.7) | 0.777<sup>M</sup> |
| **PO2 (mmHg)<sup>^</sup>** | 50.2(41.9-63.5) | 65.4(55.6-83.5) | 60.4(47.3-76) | 0.000<sup>**M</sup> |
| **PaO2/FiO2 Ratio<sup>^</sup>** | 156.8(77-233.3) | 230.8(150.3-321.1) | 195.4(121.3-287.4) | 0.000<sup>**M</sup> |
| **ARDS** |  |  |  |  |
| Not Present | 4(6.3) | 34(32.1) | 38(22.5) | 0.001<sup>*P</sup> |
| Mild | 19(30.2) | 28(26.4) | 47(27.8) |  |
| Moderate | 22(34.9) | 30(28.3) | 52(30.8) |  |
| Severe | 18(38.6)b | 14(13.2)a | 32(18.9) |  |
| **Urea<sup>^</sup> (mg/dl)** | 45(34-71) | 32(21-51) | 36(24-55) | 0.000<sup>**M</sup> |
| **Creatinine<sup>^</sup> (mg/dl)** | 1.2(1.0-1.8) | 1(0.8-1.4) | 1.1(0.9-1.5) | 0.002<sup>*M</sup> |
| **Sodium<sup>^</sup> (mg/dl)** | 136(132-139) | 137(134-139) | 136(133-139) | 0.311<sup>M</sup> |
| **Potassium<sup>^</sup>(mg/dl)** | 4.1(3.7-4.6) | 4.1(3.8-4.5) | 4.1(3.7-4.5) | 0.810<sup>M</sup> |
| **Bicarbonate<sup>^</sup> (mg/dl)** | 19(16-22) | 20(18-22) | 20(18-22) | 0.130<sup>M</sup> |
| **Total Bilirubin<sup>^</sup> (mg/dl)** | 0.6(0.4-0.9) | 0.6(0.4-0.8) | 0.6(0.4-0.9) | 0.417<sup>M</sup> |
| **SGPT<sup>^</sup> (U/L)** | 43(22-62) | 32(19-69) | 38(21-64) | 0.475<sup>M</sup> |
| **Arterial Lactate<sup>^</sup> (mmol/L)** | 2.3(1.8-3.5) | 1.7(1.3-2.3) | 1.9(1.4-3.1) | 0.001<sup>*M</sup> |
| **Serum Albumin<sup>^</sup> (g/L)** | 3.2(2.8-3.5) | 3.4(3.1-3.6) | 3.3(3.0-3.6) | 0.014<sup>*M</sup> |
| **Prothrombin Time<sup>^</sup> (sec)** | 11.4(10.7-12.5) | 11.5(10.9-12.1) | 11.5(10.8-12.5) | 0.997<sup>M</sup> |
| **APTT<sup>^</sup> (sec)** | 30.3(27.4-32.6) | 26.9(24.8-32.4) | 29.5(25.7-32.5) | 0.068<sup>M</sup> |
| **Baseline CRP^ mg/L** | 175.5(86.8-261.7) | 131.9(66.2-198.8) | 145.4(78.0-219.1) | 0.008<sup>*M</sup> |
| **CRP mg/L** |  |  |  |  |
| **≤200** | 41(61.2) | 88(75.2) | 129(70.1) | 0.046<sup>*P</sup> |
| **>200** | 26(38.8) | 29(24.8) | 55(29.9) |  |
| **Baseline D-dimer^ ng/ml** | 3729(1086-8763) | 1257.5(615-4256) | 1683(665-6858) | 0.001<sup>*M</sup> |
| **Baseline LDH^ U/L** | 555.5(452-866) | 398(312-512) | 461(334-614) | 0.000<sup>**M</sup> |
| **Baseline Ferritin^ ng/ml** | 1315(581.8-1675.6) | 668.8(337.9-1557) | 936.6(422.7-1675.6) | 0.002<sup>*M</sup> |
| **Baseline IL-6^ pg/ml** | 78.6(34.9-362) | 21.8(11.8-37.1) | 35.8(20.8-107.5) | 0.000<sup>**M</sup> |
| **Baseline Pro-calcitonin^ ng/ml** | 0.9(0.3-3.0) | 0.3(0.1-0.7) | 0.4(0.2-1.6) | 0.000<sup>**M</sup> |
| **Baseline Troponin^ ng/ml** | 18(0-154) | 6(3-20) | 9.5(4-57) | 0.000<sup>**M</sup> |
| **Blood culture** |  |  |  |  |
| Positive | 5(8.3) | 7(7.7) | 12(7.9) | 0.100<sup>f</sup> |
| Negative | 55(91.3) | 84(92.3) | 139(92.1) |  |
| **Non-invasive ventilation (in ER)** |  |  |  |  |
| Yes | 38(56.7) | 28(42.2) | 66(35.5) | 0.000<sup>**P</sup> |
| No | 29(43.3) | 91(76.8) | 120(63.2) |  |
| **CXR Findings** |  |  |  |  |
| Unilateral Radiologic Findings | 3(4.8) | 6(5.3) | 9(5.1) | 0.000<sup>**MC</sup> |
| Bilateral Radiologic Findings | 59(93.7)a | 88(77.2) | 147(83.1) |  |
| Multilobar Infiltrates | 36(57.1)a | 30(26.3) | 66(37.3) |  |
| Consolidation | 32(50.8)a | 32(28.1) | 64(36.2) |  |
| Pleural Effusion | 3(4.8) | 7(6.1) | 10(5.6) |  |
| Others | 7(11.1) | 10(8.8) | 17(9.6) |  |
| Normal | 0 | 16(14.0) | 16(9.0) |  |
| **Clinical Severity Scores** |  |  |  |  |
| **SOFA score^** | 6(5-8) | 4(4-6) | 5(4-8) | 0.000<sup>**M</sup> |
| **CURB-65 score^** | 2(1-3) | 1(0-2) | 1(1-2) | 0.000<sup>**M</sup> |
| **Mulbsta^** | 7(5-9) | 4(0-7) | 5.5(2-8) | 0.000<sup>*M</sup> |
| <sup>^</sup>Median (IQR), <sup>^^</sup>Mean±SD, <sup>*</sup>P-value<0.05, <sup>**</sup>P-value<0.0001, <sup>T</sup>: Independent sample T test, <sup>M</sup>: Mann Witney U test,<br><sup>P</sup>: Pearson Chi-Square, <sup>f</sup>: Fisher's Exact Test, <sup>MC</sup>: Chi-square for multiple Responses |  |  |  |  |

There was no difference in the arterial pH or arterial PCO2 but arterial pO2 was lower among non survivors (50.2 vs 65.4 mm Hg, p<0.001). PaO2/FiO2 ratio was lower among non-survivors (156.8 vs 230.8, p<0.001). Serum creatinine and arterial lactate were higher amongst non-survivors (1.2 vs 1.0 mg/dl, p=0.002; 2.3 vs 1.7 mmol/L, p=0.001 respectively). Similarly, CRP (175.5 vs 131.9 mg/L, p=0.008), LDH (555.5 vs 398 U/L, p<0.001), d-Dimer (3729 vs 1257 ng/ml, p=0.001),Ferritin (1315 vs 668.8 ng/ml, p=0.002), IL-6 (78.6 vs 21.8 pg/ml, p<0.001), pro-calcitonin (0.9 vs 0.3 ng/ml, p<0.001) and Troponin-I (18 vs 6 ng/ml, p<0.001) were all higher amongst non-survivors compared to survivors. Non-survivors in our cohort had higher SOFA score, CURB-65 score and Mulbsta scores (6 vs 4, 2 vs 1, 7 vs 4 respectively, p<0.001 for all). Severe ARDS was more prevalent (38.6% vs 13.2%, p=0.001) among non survivors compared to survivors.

### Hospital Course and outcome

ICU admission was more common among non-survivors (93.8% vs 34.2%) (Table 3). Hence, invasive ventilation was more frequent among non-survivors too (86.6% vs 6.7%). Most common in-hospital complication was Acute Kidney Injury (58.2% vs 7.6%) in non-survivors.

**Table 3:**
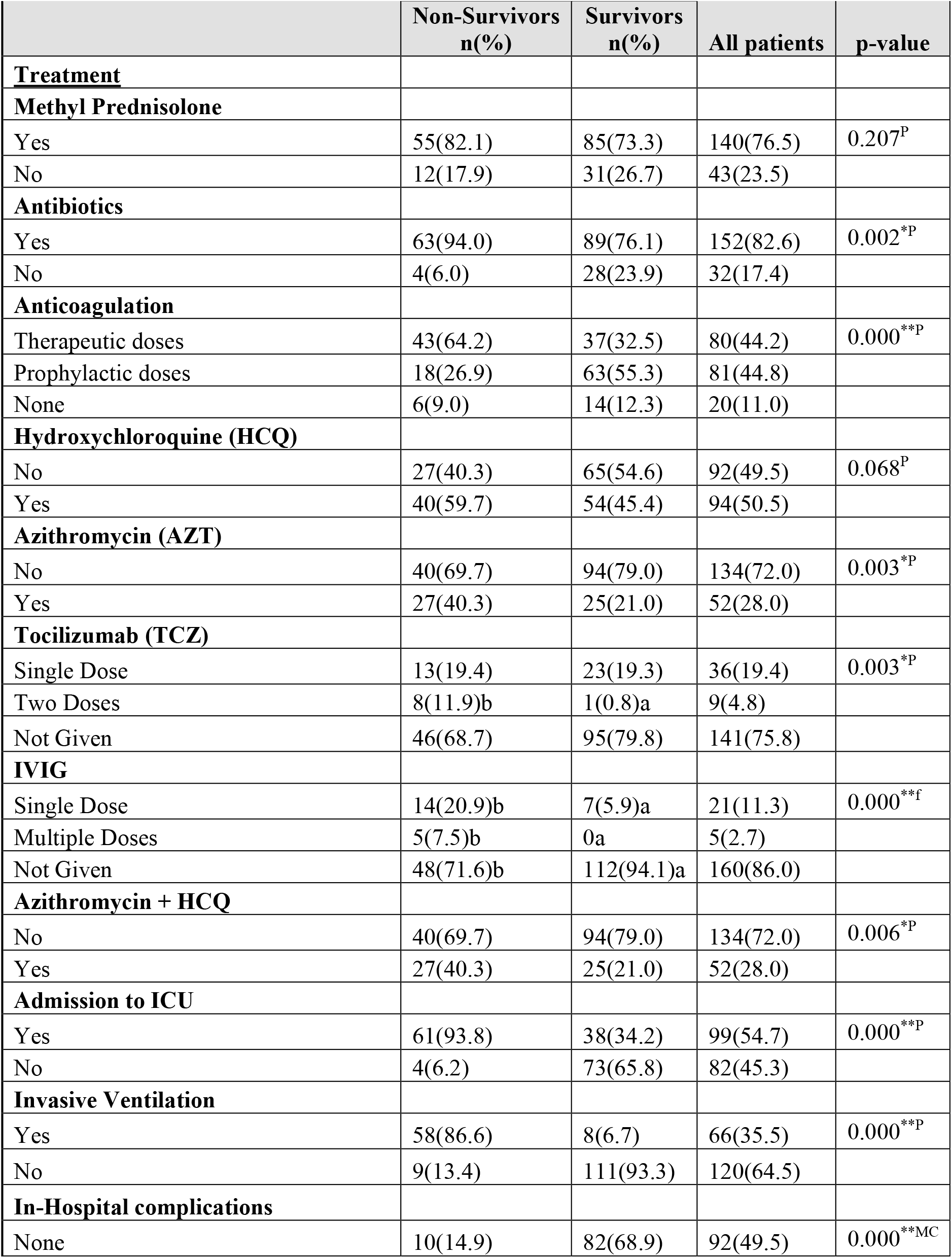
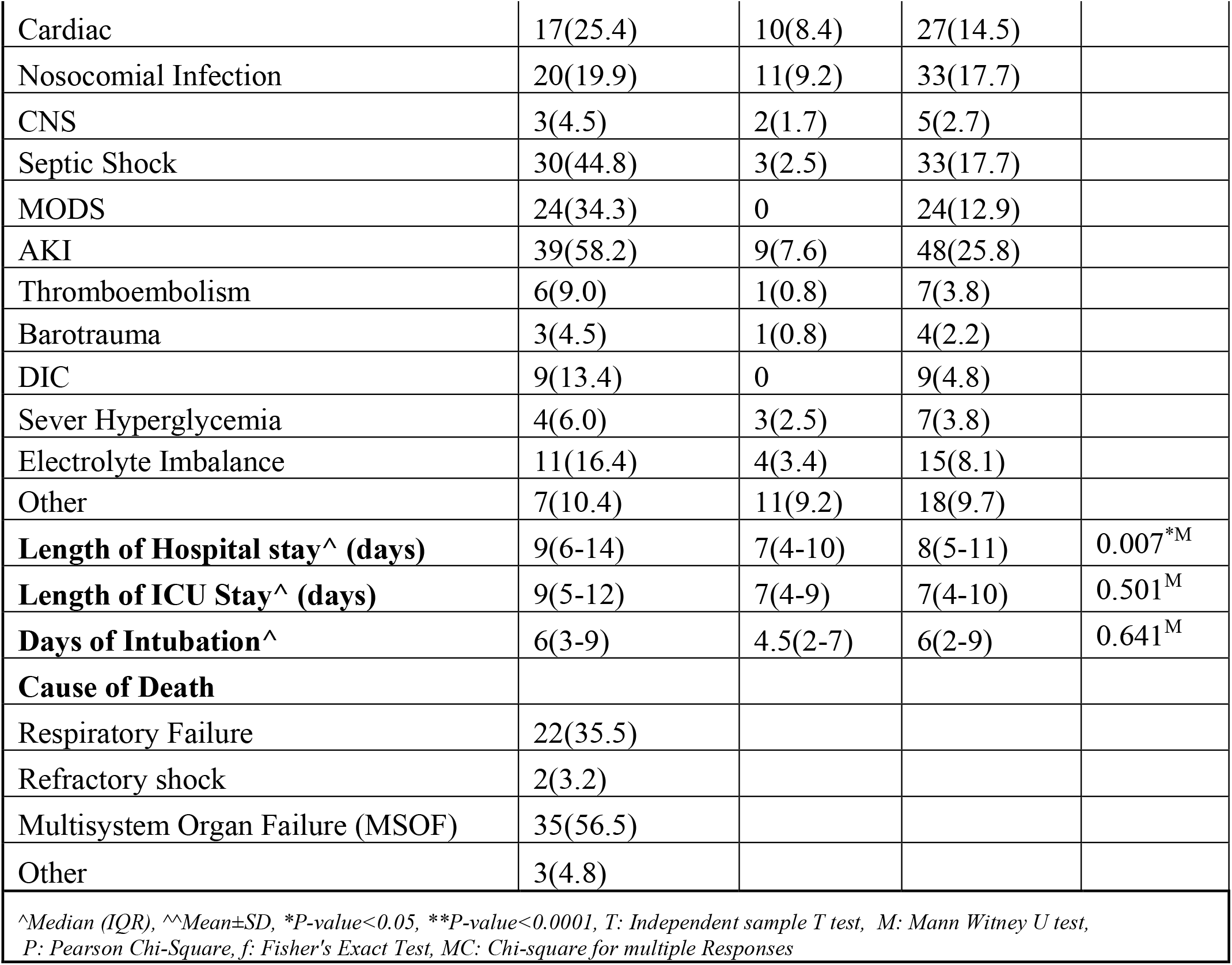
Hospital course by mortality.

| Table 3: Hospital course by mortality |  |  |  |  |
| --- | --- | --- | --- | --- |
|  | Non-Survivors<br>n(%) | Survivors<br>n(%) | All patients | p-value |
| **Treatment** |  |  |  |  |
| **Methyl Prednisolone** |  |  |  |  |
| Yes | 55(82.1) | 85(73.3) | 140(76.5) | 0.207<sup>P</sup> |
| No | 12(17.9) | 31(26.7) | 43(23.5) |  |
| **Antibiotics** |  |  |  |  |
| Yes | 63(94.0) | 89(76.1) | 152(82.6) | 0.002<sup>*P</sup> |
| No | 4(6.0) | 28(23.9) | 32(17.4) |  |
| **Anticoagulation** |  |  |  |  |
| Therapeutic doses | 43(64.2) | 37(32.5) | 80(44.2) | 0.000<sup>**P</sup> |
| Prophylactic doses | 18(26.9) | 63(55.3) | 81(44.8) |  |
| None | 6(9.0) | 14(12.3) | 20(11.0) |  |
| **Hydroxychloroquine (HCQ)** |  |  |  |  |
| No | 27(40.3) | 65(54.6) | 92(49.5) | 0.068<sup>P</sup> |
| Yes | 40(59.7) | 54(45.4) | 94(50.5) |  |
| **Azithromycin (AZT)** |  |  |  |  |
| No | 40(69.7) | 94(79.0) | 134(72.0) | 0.003<sup>*P</sup> |
| Yes | 27(40.3) | 25(21.0) | 52(28.0) |  |
| **Tocilizumab (TCZ)** |  |  |  |  |
| Single Dose | 13(19.4) | 23(19.3) | 36(19.4) | 0.003<sup>*P</sup> |
| Two Doses | 8(11.9)b | 1(0.8)a | 9(4.8) |  |
| Not Given | 46(68.7) | 95(79.8) | 141(75.8) |  |
| **IVIg** |  |  |  |  |
| Single Dose | 14(20.9)b | 7(5.9)a | 21(11.3) | 0.000<sup>**f</sup> |
| Multiple Doses | 5(7.5)b | 0a | 5(2.7) |  |
| Not Given | 48(71.6)b | 112(94.1)a | 160(86.0) |  |
| **Azithromycin + HCQ** |  |  |  |  |
| No | 40(69.7) | 94(79.0) | 134(72.0) | 0.006<sup>*P</sup> |
| Yes | 27(40.3) | 25(21.0) | 52(28.0) |  |
| **Admission to ICU** |  |  |  |  |
| Yes | 61(93.8) | 38(34.2) | 99(54.7) | 0.000<sup>**P</sup> |
| No | 4(6.2) | 73(65.8) | 82(45.3) |  |
| **Invasive Ventilation** |  |  |  |  |
| Yes | 58(86.6) | 8(6.7) | 66(35.5) | 0.000<sup>**P</sup> |
| No | 9(13.4) | 111(93.3) | 120(64.5) |  |
| **In-Hospital complications** |  |  |  |  |
| None | 10(14.9) | 82(68.9) | 92(49.5) | 0.000<sup>**MC</sup> |
| Cardiac | 17(25.4) | 10(8.4) | 27(14.5) |  |
| Nosocomial Infection | 20(19.9) | 11(9.2) | 33(17.7) |  |
| CNS | 3(4.5) | 2(1.7) | 5(2.7) |  |
| Septic Shock | 30(44.8) | 3(2.5) | 33(17.7) |  |
| MODS | 24(34.3) | 0 | 24(12.9) |  |
| AKI | 39(58.2) | 9(7.6) | 48(25.8) |  |
| Thromboembolism | 6(9.0) | 1(0.8) | 7(3.8) |  |
| Barotrauma | 3(4.5) | 1(0.8) | 4(2.2) |  |
| DIC | 9(13.4) | 0 | 9(4.8) |  |
| Sever Hyperglycemia | 4(6.0) | 3(2.5) | 7(3.8) |  |
| Electrolyte Imbalance | 11(16.4) | 4(3.4) | 15(8.1) |  |
| Other | 7(10.4) | 11(9.2) | 18(9.7) |  |
| **Length of Hospital stay^ (days)** | 9(6-14) | 7(4-10) | 8(5-11) | 0.007<sup>*M</sup> |
| **Length of ICU Stay^ (days)** | 9(5-12) | 7(4-9) | 7(4-10) | 0.501<sup>M</sup> |
| **Days of Intubation^** | 6(3-9) | 4.5(2-7) | 6(2-9) | 0.641<sup>M</sup> |
| **Cause of Death** |  |  |  |  |
| Respiratory Failure | 22(35.5) |  |  |  |
| Refractory shock | 2(3.2) |  |  |  |
| Multisystem Organ Failure (MSOF) | 35(56.5) |  |  |  |
| Other | 3(4.8) |  |  |  |
| <sup>^</sup>Median (IQR), <sup>^^</sup>Mean±SD, *P-value<0.05, **P-value<0.0001, T: Independent sample T test, M: Mann Witney U test,<br>P: Pearson Chi-Square, f: Fisher's Exact Test, MC: Chi-square for multiple Responses |  |  |  |  |

Use of antibiotics (82% vs 73%, p=0.002), Hydroxychloroquine (HCQ) (59.7% vs 45.4%, p=0.06), Azithromycin (AZT) (55% vs 32.8%, p=0.003), combination of HCQ and AZT (40% vs 21%, p=0.006) and intravenous immunoglobulin (IVIG) (p<0.001) were all associated with mortality in our cohort. Use of therapeutic anticoagulation was associated with mortality (p<0.001). Single dose of Tocilizumab showed no mortality benefit while two doses were associated with mortality (p=0.003). There was no mortality benefit of using methylprednisolone in our patients (p>0.05).

Length of hospital stay was longer in non-survivors (9 days vs 7 days, p=0.007). Most common in-hospital complication among non-survivors compared to survivors was Acute kidney injury followed by septic shock (p<0.001). Most common cause of death was multisystem organ failure (35/67, 56.5%).

### Predictors of in-hospital all-cause mortality

Univariable binary logistic regression (Table 4) at a cut-off of p<0.10, showed that age greater than 60 years, diabetes mellitus, unit increase in respiratory rate, temperature, SOFA, CURB-65, Mulbsta scores, oxygen saturation < 93%, NLR greater than 5, thrombocytopenia, multilobar involvement on chest x-ray, unit increase in arterial lactate, CRP higher than 200 mg/L, pro-calcitonin higher than 2 ng/ml, high D-dimer, Ferritin and IL-6 showed high odds of mortality. ICU admission, use of antibiotics, therapeutic anticoagulation, Tocilizumab, IVIG, non-invasive ventilation in emergency room, unit increase in length of stay (days), electrolyte abnormalities, sepsis, cardiac complications and acute kidney injury were associated with high odds of mortality. While unit decrease in lymphocyte count, arterial pO2, PaO2/FiO2 ratio and serum albumin were related to high odds of mortality.

**Table 4:** Univariable binary logistic regression for predictors of in-hospital all-cause mortality.

|  |  |  |  |
| --- | --- | --- | --- |
|  | Crude OR | 95% CI | p-value |
| Age >60 years | 2.94 | 1.58 - 5.48 | 0.001 |
| Gender (male) | 1.06 | .52- 2.18 | 0.859 |
| Diabetes mellitus | 1.86 | 1.02- 3.42058 | 0.04 |
| Respiratory rate /min | 1.08 | 1.04- 1.12 | 0.000 |
| Temperature °F | 1.50 | 1.14- 2.09 | 0.006 |
| Oxygen saturation <93% | 4.51 | 2.23- 9.14 | 0.000 |
| SOFA | 1.35 | 1.13- 1.61 | 0.001 |
| CURB-65 | 1.96 | 1.43- 2.71 | 0.000 |
| Mulbsta | 1.24 | 1.14- 1.37 | 0.000 |
| White blood cells (x10E9/L) | 1.05 | .99- 1.10 | 0.068 |
| Neutrophils (%) | 1.07 | 1.04- 1.12 | 0.000 |
| Lymphocytes (%) | 0.91 | .87- .96 | 0.001 |
| NLR>=5 | 3.64 | 1.65- 8.08 | 0.001 |
| Platelets (x10E9/L) | 0.99 | .99- 1.00 | 0.076 |
| Thrombocytopenia (yes) | 11.6 | 1.37- 98.59 | 0.025 |
| Lymphopenia (yes) | 1.76 | .96- 3.24 | 0.066 |
| Multilobar involvement on CXR | 3.44 | 1.83- 6.49 | 0.000 |
| arterial pH | 1.49 | .06- 37.70 | 0.807 |
| arterial PO2 mmHg | 0.96 | .94- .98 | 0.000 |
| Pao2/Fio2 ratio | 0.51 | .37- .71 | 0.000 |
| Creatinine mg/dl | 0.96 | .86- 1.08 | 0.54 |
| Arterial lactate (mmol/L) | 1.51 | 1.11- 2.05 | 0.009 |
| Albumin g/L | 0.34 | .16- .72 | 0.005 |
| CRP(>200) mg/L | 1.92 | 1.01- 3.67 | 0.047 |
| LDH U/L | 1.00 | .99- 1.00 | 0.215 |
| D-dimer ng/ml | 1.00 | .99- 1.00 | 0.054 |
| Ferritin ng/ml | 1.00 | 1.00- 1.00 | 0.017 |
| IL-6 pg/ml | 0.99 | .99- 1.00 | 0.048 |
| Pro-calcitonin >2 ng/ml | 3.23 | 1.48- 7.07 | 0.003 |
| Troponin ng/ml | 1.00 | .99- 1.00 | 0.276 |
| Antibiotics | 4.95 | 1.65- 14.82 | 0.004 |
| **Anticoagulation** |  |  |  |
| No Anticoagulation | 1 |  |  |
| Therapeutic anticoagulation | 2.7 | 0.94-7.76 | 0.074 |
| Prophylactic anticoagulation | 0.66 | 0.224-1.98 |  |
| **Use of Tocilizumab** |  |  |  |

| No | 1 |  |  |
| --- | --- | --- | --- |
| Single dose | 1.17 | 0.54-2.51 | 0.002 |
| Double dose | 16.5 | 2.01- 136.06 |  |
| IVIG | 4.66 | 1.77- 12.29 | 0.002 |
| ICU admission | 29.29 | 9.90- 86.69 | 0.000 |
| NIV in Emergency room | 4.25 | 2.24-8.10 | 0.000 |
| Invasive Ventilation | 89.4 | 32.76- 244.01 | 0.000 |
| Length of hospital stay (days) | 1.03 | .99- 1.08 | 0.114 |
| Electrolyte abnormalities | 5.6 | 1.72- 18.53 | 0.004 |
| Sepsis | 31.35 | 9.04-108.68 | 0.000 |
| Cardiac complications | 3.7 | 1.58- 8.67 | 0.003 |
| Acute Kidney injury | 17.0 | 7.3- 39.24 | 0.000 |

Multivariable binary logistic regression (Fig. 2) using all the above variables in order of significance at a p-value <0.05 showed that age more than 60 years (OR=4.27; 95% CI=1.08-16.8); Oxygen saturation <93% (OR=11.56; 95% CI=1.2-105.9) serum prolactin level higher than 2 ng/ml (OR=31.5;95% CI=3.4-288.2); unit increase in SOFA score (OR=1.5; 95% CI=1.02-2.23); unit increase in body temperature (OR=2.17; 95% CI= 1.07-4.39); ICU admission (OR=16.9; 95% CI=2.8-100.5) and sepsis (OR=23.6; 95% CI=4.6-119.9) were associated with high odds of mortality.

**Figure 2:**
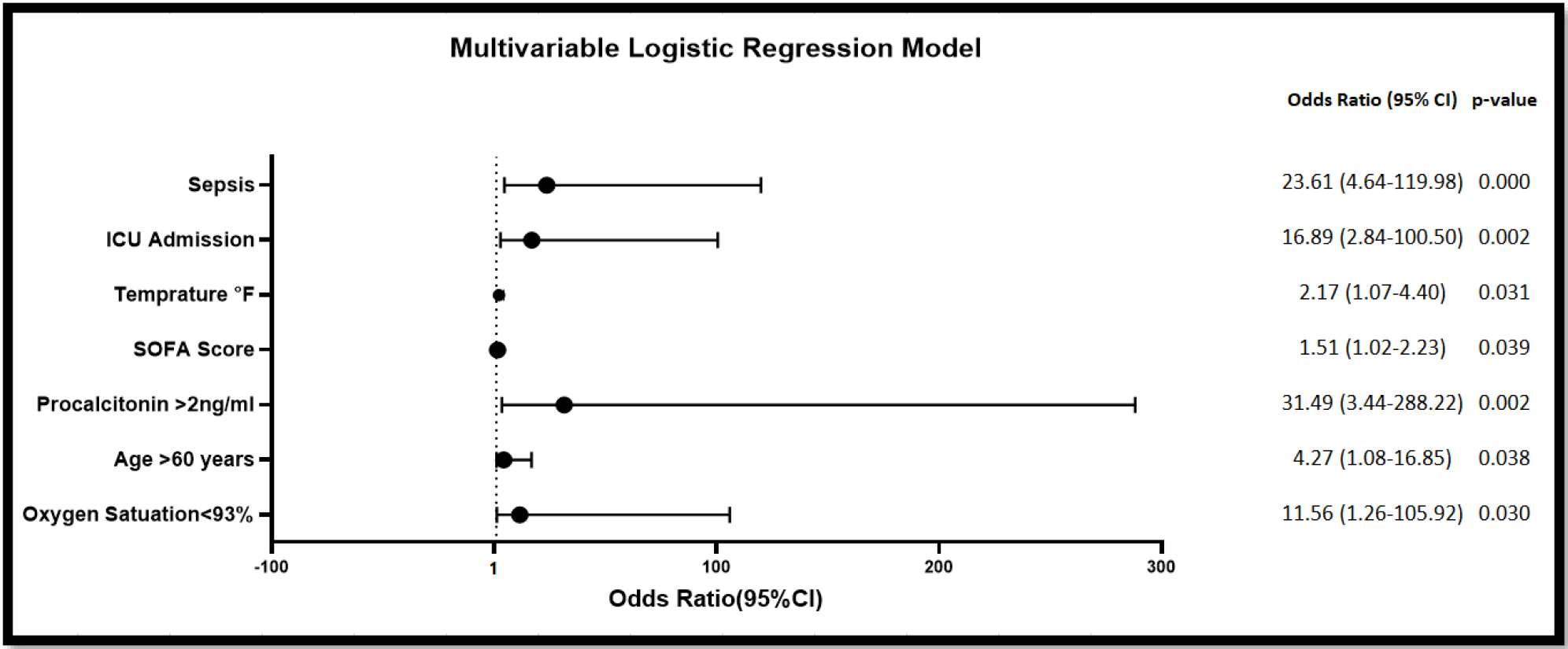
Multivariable Logistic regression model of all-cause mortality in COVID-19 patients.

## Discussion

In this prospective cohort study, we report the clinical attributes and risk factors associated with all-cause mortality among hospitalized COVID-19 patients. We found the all-cause mortality to be 36% which is disproportionately high (88%) in those who were ventilated. It is important to highlight here that our patient pool had more critical patients (approximately 74% of total admitted) with 31% and 19% having moderate and severe ARDS respectively on presentation while 35% needing non-invasive ventilation (NIV) in ER to manage respiratory failure. ICU admission was necessary for 55% of which 35% required mechanical ventilation (MV). Our patient population appears to be sicker compared to the only other unpublished data from the city. The reason for this difference could be admission of non-critical patients for monitoring and isolation since it was a private hospital [6] On the contrary, as mentioned before, our center is a philanthropic, free of cost referral centre for the underprivileged with limited bed capacity. Hence, admission was strictly reserved for critical patients. The poor survival in ventilated cases, apart from the natural disease process, may be due to poor knowledge of the pathogenic mechanism of respiratory injury in the initial days. Most patients were managed with early ventilator support to avoid fatigue and potential risk of aerosolization of COVID-19 with NIV [7]. Gradual understanding of the disease process has shifted the management strategy from early mechanical ventilation towards use of NIV till tolerated as suggested by the National and International COVID guidelines [3, 5]. Global mortality from COVID-19 varies widely (20% -97%) [8, 9] depending on ICU facilities, ventilator performance, experience of ICU team, patient and disease characteristics, geographic area, seasonality and duration of follow up [4]. High ventilator mortality has been reported even from the best centers in Wuhan (97%), New York (88%), UK (67%) and Italy (53.4%) [10-13] questioning the role of invasive ventilation in COVID-19 management especially in LMIC [14]. Ventilator induced lung injury due to barotrauma, volutrauma, atelectrauma, oxytrauma and infections further jeopardize the outcome of COVID-19 patients [15]. Hospitalized patients with COVID-19 have 5 times higher reported mortality than those with influenza pneumonia [16].

Non-survivors in our study showed worse clinical profile with low oxygen saturation, higher temperature and respiratory rate along with bilateral, multi-lobar infiltrates on chest x-ray and raised inflammatory parameters at presentation. This indicates that they were already in the late phases of CRS at admission [17]. The median time to hospitalization from onset of symptoms was 7 days for both survivors and non-survivors consistent with reported literature [18]. However, why some patients were more prone to develop CRS by day 7 is not clear. Older age and male sex were found to be associated with higher mortality as reported globally [19, 20]. It is important to note that the median age of our overall COVID-19 cohort (3855 patients) was 20 years younger than this subset who got admitted to the isolation unit. This is consistent with the idea that since Pakistan’s age composition is largely of younger age group, most people who are infected with COVID-19 go asymptomatic or have mild symptoms [21]. Among the high risk comorbid conditions for COVID-19 listed by CDC, only diabetes was statistically significant in our cohort (crude OR= 1.86;95%CI=1.02-3.42)[22].

Neutrophillia, lymphopenia and thrombocytopenia were more pronounced among non-survivors with NLR > 5 having an odds of 3.64 (95%CI=1.65-8.08). Wu C et al found a significant association between neutrophillia, lymphopenia (peripheral CD3, CD4, and CD8 T-cell counts decreased) and development of ARDS [23]. As observed in literature, CRP, D-dimer and IL-6 were all higher among non-survivors in our cohort [23]. Although baseline pro-calcitonin was low in the cohort overall (median 0.4 ng/ml), value of greater than 2 ng/ml was strongly associated with mortality. It is uncertain whether this suggests secondary bacterial infection or hyper-inflammation as studies have suggested that raised pro-calcitonin as a marker of bacterial infection tends to lose specificity as COVID progresses [20, 23]. Non-survivors in our cohort developed acute kidney injury, sepsis and multi organ dysfunction syndrome (MODS) more frequently than survivors. Unit increase in SOFA score was associated with high odds of mortality in our data. Since SOFA tends to reflect the effect on multiple organ systems, it has proved to be a better predictor of mortality in COVID-19 [24].

Some desperate therapies used globally including Hydroxychloroquine and azithromycin (alone or in combination), therapeutic anticoagulation, antibiotics, two doses of Tocilizumab and IVIG showed increased odds of mortality which when adjusted for other factors became insignificant in the final model. HCQ despite inhibiting viral replication in vitro [25] did not prove beneficial in RCTs [26], rather proved to be toxic. This led to the FDA revoking its emergency use authorization in June 2020 [27]. AZT is the only effective oral drug for treatment of XDR salmonella [28] and COVID-19 pandemic has led to its mass injudicious use, both over the counter and prescription driven, which may increase antimicrobial resistance in the long run. Studies have failed to demonstrate any benefit of AZT alone [29] or in combination with HCQ [30] with a risk of compounding cardiac toxicity by QTc prolongation [31]. Antibiotics were started in 82.6% of our patients suspecting respiratory bacterial co-infection although only 7% initial blood cultures were positive (most patients were not producing sputum) and markers like WBC count and pro-calcitonin were not elevated in the majority. Literature reports low rates of secondary infection with COVID-19 (only 8% in a review of 9 studies) with paradoxical high consumption (72%) of broad spectrum antibiotics [32]. We believe COVID-19 specific antibiotic stewardship guidance is essential to stop the rampant over-use of antibiotics especially in LMIC countries like Pakistan where AMR is already high. Steroids have shown benefit in severe and critical COVID-19 in the RECOVERY trial [33] and a recent meta-analysis of 7 trials conducted on 1703 patients showed a reduction in 28-day mortality compared with standard care or placebo (32% vs 40%, OR= 0.66,95%CI=0.53-0.82)[34]. However, most experience is with dexamethasone and not methyl prednisolone although MP has been recommended as an alternate to dexamethasone in a dose of 32mg/d in current guidelines [35]. Our patient population was given MP but in higher doses (40mg q8hrs) which may have resulted in the observed hyperglycemia, secondary infections and electrolyte imbalance in our data. Moreover, a randomized trial on severe COVID-19 patients in Brazil did not show any mortality benefit of MP at 28 days as compared to placebo (37% vs 38%)[36]. Some observational studies have reported mortality benefit of TCZ [37, 38] but RCTs failed to demonstrate any difference in survival when compared to placebo or usual care [39]. Data from our center also did not show any survival benefit of TCZ [40].

The final model showed that the factors associated with mortality are older age, low oxygen saturation, high SOFA scores, raised pro-calcitonin, ICU admission and sepsis. This is the first prospective cohort study from Pakistan on in-hospital mortality of COVID-19 patients. Detailed clinical history, laboratory parameters and therapeutics have been compared. There is no attrition as patients were followed till discharge or death. Despite the limitations of a small sample we were able to detect many predictors of mortality. The study is limited by data from a single-center with critically ill COVID-19 patients which may introduce a selection bias and inflate the mortality. Hence, results from this study may help in the risk stratification and management of similar critically ill patients only. The sample size is limited because of the number of beds available at the unit during the first wave. We plan to add more numbers in future as we are currently seeing the second wave. A larger multi-center cohort study from various hospitals of the country would help to further validate the findings of our study.

## Supporting information

IRB statement of compliance

IRB apporval

Strobe checklist

## Data Availability

available on request

