## Supplementary material for "Determinants of in-hospital mortality in COVID-19; a prospective cohort study from Pakistan": IRB statement of compliance

IRD Global Limited
15 Beach Road # 02-01
Singapore 189677.
T: +65 6372 8778
F: +65 6372 8777
www.ird.global

**Interactive Research & Development IRB (IRD-IRB)**
**Statement of Compliance &**
**Designated IRB status for The Indus Hospital**

Interactive Research & Development (IRD) is an international grants-based research and service organization working in the health and development sector. IRD's independent institutional review board (IRD-IRB) conducts review in accordance with pertinent authorities, including but not limited to, the International Conference on Harmonization (ICH) Guidelines for Good Clinical Practice E6, United States Food and Drug Administration (21 CFR Parts 50 and 56), U.S. Department of Health and Human Services (45 CFR Part 46), the ethical principles outlined in the Belmont Report, and the Council for International Organization of Medical Sciences (CIOMS) international ethical guidelines 2002.

IRD-IRB's mandate is to protect the rights and welfare of individuals who volunteer to participate in research managed by IRD, in research where IRD is either a partner or a collaborator or for institutes that have asked IRD to partner them in their institutional research.

IRD-IRB is appropriately constituted, organized, and operated in accordance with regulations and guidelines referenced above and the World Medical Association Declaration of Helsinki, to the extent applicable.

Interactive Research & Development (IORG0004336) is registered with the U.S. Department of Health and Human Services (DHHS) Office for Human Research Protections organization with an IRB# 00005148 (effective through Nov 19, 2023). IRD-IRB has federal wide assurance (FWA) for the protection of Human Subjects for International (non US) Institutions FWA00023738 expiration date Dec 18, 2025. In additions, IRD-IRB is the designated IRB for The Indus Hospital (FWA # 00016337) for the review and continuing oversight for all its human subject research vide an authorization agreement with IRD (Expires Dec 16, 2025).

In addition, IRD-IRB utilizes electronic signatures compliant with 21 CFR Part 11.

Salma H. Badruddin, PhD
Chair – IRD Institutional Review Board
