## Supplementary material for "Determinants of in-hospital mortality in COVID-19; a prospective cohort study from Pakistan": IRB apporval

**IRD Global Limited**
15 Beach Road # 02-01
Singapore 189677.
T: +65 6372 8778
F: +65 6372 8777
www.ird.global

### Protection of Human Subjects – Declaration / Assurance of IRB Approval

| **PI** | **IRD-IRB #** | **Department/Institute** |
|---|---|---|
| Samreen Sarfaraz | IRD_IRB_2020_04_002 | ID/TIH |
| **Approval Date** | **Expiration Date** | **Administrative Due Date** |
| 09-Apr-2020 | N/A | N/A |

**The following research study has been reviewed by the IRD-IRB:**
Epidemiology, geographical distribution and outcome of patients infected with SARS-CoV-2 at The Indus Hospital, Karachi, Pakistan

**IRB EXPEDITED STATUS: APPROVED**
The IRD-IRB has reviewed the above-referenced study and determined that, as currently described, it was eligible for expedited review and has been approved, as per the following category:

**Category #5: Research involving materials (data, documents, records, or specimens) that have been collected, or will be collected solely for nonresearch purposes (such as medical treatment or diagnosis).**

Stamped consent form(s) [if applicable] are attached for your reference.

As principal investigator for a study involving human subjects, you assume certain responsibilities, specifically:

1. You will conduct the study according to the protocol approved by the IRB. As the PI, you will be accountable for your own research and the protection of human subjects. You will ensure, at all times, that you have the appropriate resources and facilities to conduct the study. You will ensure that all research personnel involved in the conduct of the study have been appropriately trained on the protection of human subjects, in addition to the study procedures.

2. Any unanticipated problems involving risks to participants or others will be reported to the IRB in accordance to the IRB policy. Changes in approved research initiated without IRB approval to eliminate apparent immediate hazards to the participant, are to be reported to the IRB.

3. Any changes in your research plan must be submitted to the IRB for review and approval prior to implementation of the change. Proposed changes in approved research cannot be initiated without IRB approval, except when necessary to eliminate apparent immediate hazards to participants.

4. **A Progress Report for continuing review must be submitted to the IRB administration by the administrative due date in order to allow sufficient time for review to be completed prior to the expiration date.** Failure to attain continued renewal by the expiration date will result in the study being assigned an *inactive* status, whereby all research activities including data analysis must stop immediately.

| Signature of the IRB Chair/Designee | |
|---|---|
| Name / Designation<br>Dr Salma H. Badruddin/ Chair, IRD-IRB | Date: 09-Apr-2020 |
